# CORRELATION BETWEEN CHEST COMPUTED TOMOGRAPHY AND LUNG ULTRASONOGRAPHY IN PATIENTS WITH CORONAVIRUS DISEASE 2019 (COVID-19)

**DOI:** 10.1101/2020.05.08.20095117

**Authors:** Yale Tung-Chen, Milagros Martí de Gracia, Aurea Díez-Tascón, Sergio Agudo-Fernández, Rodrigo Alonso-González, Pablo Rodríguez-Fuertes, Luz Parra-Gordo, Silvia Ossaba-Vélez, Rafael Llamas-Fuentes

**Affiliations:** Department of Emergency Medicine, Hospital Universitario La Paz (Madrid, Spain).; Department of Medicine, Universidad Alfonso X El Sabio (Madrid, Spain); Department of Emergency Radiology, Hospital Universitario de La Paz, Madrid, Spain; Department of Emergency Medicine, Hospital Universitario Reina Sofía (Cordoba, Spain).

**Keywords:** Coronavirus Disease 2019 (COVID-19), Severe acute respiratory syndrome coronavirus 2 (SARS-CoV-2), Lung ultrasonography (LUS), chest Computed Tomography (chest CT).

## Abstract

**INTRODUCTION:** There is growing evidence regarding the imaging findings of Coronavirus Disease 2019 (COVID-19), in chest X-ray and Computed Tomography scan (CT). At this moment, the role of Lung Ultrasonography (LUS) has yet to be explored.

**OBJECTIVES:** The main purpose of this study is to evaluate the correlation between LUS findings and chest CT in confirmed (positive RT-PCR) or clinically highly suspicious (dyspnea, fever, myasthenia, gastrointestinal symptoms, dry cough, ageusia or anosmia) of COVID-19 patients.

**METHODS:** Prospective study carried out in the emergency department (ED) of confirmed or clinically highly suspicious COVID-19 patients who were subjected to a chest CT and concurrent LUS exam. An experienced ED physician performed the LUS exam blind to the clinical history and results of the CT scan, which were reviewed by two radiologists in consensus for signs compatible with COVID-19 (bilateral ground-glass opacities in peripheral distribution). Compatible LUS exam was considered a bilateral pattern of B-lines, irregular pleural line and subpleural consolidations.

**RESULTS:** Between March and April 2020, fifty-one patients were consecutively enrolled. The indication for CT was a negative or indeterminate RT-PCR test (49.0%) followed by suspicion of pulmonary embolism (41.2%). Radiological signs compatible with COVID-19 were present in thirty-seven patients (72.5%) on CT scan and forty patients (78.4%) on LUS exam. The presence of LUS findings was correlated with a positive CT scan suggestive of COVID-19 (OR: 13.3, 95%CI: 4.5-39.6, *p*<0.001) with a sensitivity of 100.0% and a specificity of 78.6%, positive predictive value of 92.5% and negative of 100.0%. There was no missed diagnosis of COVID-19 with LUS compared to CT in our cohort. The LUS Score had a good correlation with CT total severity score (ICC 0.803, 95% CI 0.60-0.90, *p*<0.001).

**CONCLUSION:** LUS presents similar accuracy compared to chest CT to detect lung abnormalities in COVID-19 patients.

**Summary Statement:** LUS presents similar accuracy compared to chest CT to detect lung abnormalities in COVID-19 patients.

**Key Results:**

- Common LUS findings mirror those previously described for CT: bilateral, peripheral, consolidation and/or ground glass opacities.
- LUS findings correlated with CT scan with a sensitivity of 100.0% and a specificity of 78.6%, positive predictive value of 92.5% and negative of 100.0%. The Lung score had a good correlation with CT total severity score (ICC 0.803, 95% CI 0.601-0.903, *p* < 0.001).
- There were no missed diagnosis of COVID-19 with LUS compared to CT in our cohort.

## INTRODUCTION

Coronavirus Disease 2019 (COVID-19) is a highly contagious illness caused by the Severe Acute Respiratory Syndrome Coronavirus 2 (SARS-CoV-2). The 11^th^ of March of 2020, the World Health Organization declared a pandemic caused by SARS-CoV-2, with the spread to more than 180 countries, 2.954.106 cases confirmed and 205.398 deaths caused (1).

In this emergency, is critical the ability to quickly confirm and characterize a suspected case, moreover as almost any emergency department (ED) will struggle to keep up with the increasing number of patients and the shortage of health resources. The main diagnostic method is the reverse transcription polymerase chain reaction (RT-PCR) of the nucleic acid of SARS-CoV-2 has many limitations such as the low sensitivity or the technical difficulties to perform it (2).

There is growing evidence regarding the imaging findings of COVID-19. The most common form of radiographic presentation is the presence of a local or bilateral patchy shadowing infiltrate on chest X-ray, although with low sensitivity (absent in more than 40% of the cases) (3). CT scan shows with higher sensitivity ground glass opacities (GGO) (4), being the reason why it has been proposed as the main imaging test, incorporated in different therapeutic and triage strategies since the outbreak started (5).

The use of chest CT remains very limited due to some notable drawbacks. For mild illness, radiation exposure and overuse of health care resources, or lack thereof ability to get a CT scan seems to overshadow the need. In the critically ill, the transport of unstable patients and exposure of infected patients may also outweigh the clinical benefit. Therefore, we need alternative modalities to quickly characterize our patients.

Ultrasound machines are widely available and therefore Lung Ultrasonography (LUS) can be performed in few minutes, in mild or even unstable patients, at different hospital settings (6). Although there is an ongoing debate about how it should be applied, there is a general consensus about its usefulness (7-8). In this pandemic, the presence of subpleural consolidations, irregular pleural line and B-lines are highly suggestive for COVID-19 pneumonia (8-9).

In this Global Public Health Emergency, the evidence about the role of this technique with comparison to chest CT is limited, and needs to be further defined, in order to minimize the infectious risks.

## PATIENTS AND METHODS

Prospective study carried out in the emergency department (ED) of an academic hospital in Spain. The study was conducted in accordance with the Declaration of Helsinki, and was approved by the Research Ethics Committee of our University Hospital (PI-4089). Informed consent was obtained from each enrolled patient.

### Patient selection

Patients admitted to the ED with RT-PCR proven COVID-19, negative or indeterminate RT-PCR but clinically highly suspicious COVID-19 (dyspnea, fever, myasthenia, gastrointestinal symptoms, dry cough, ageusia or anosmia) that required a chest CT for evaluation.

The main indication for CT was a negative or indeterminate RT-PCR but clinically highly suspicious of COVID-19 or suspicion of pulmonary embolism (PE).

We excluded patients <18 years or who refused to participate. A convenience sample of fifty-one patients who met these inclusion criteria were consecutively enrolled and prospectively studied.

### Initial patient assessment

Initial evaluation of the patients included recording medical history: demographic data, comorbidities, symptoms; physical exam: temperature, blood pressure, heart rate, respiratory rate and oxygen saturation; laboratory tests: hemogram, basic metabolic panel (glucose, electrolytes, kidney function, liver enzymes, etc.), lactate dehydrogenase (LDH), ferritin, interleukin-6 (IL-6), C-reactive protein (CRP), procalcitonin and coagulation (D-dimer, INR, PTT, Fibrinogen).

### Chest CT data collection

Non-contrast chest CT scans were obtained using a multi-detector CT: SOMATOM go.Up (Siemens Healthliners). Scanning was performed with patient in supine position, and at end inspiration. The scans were acquired and reconstructed as axial images using the following parameters: 1.5 mm section thickness, 0.7 mm interval, 130 kVp. Low dose protocol was implemented with an average CTDIvol of 2 mGy Our routine protocol for patients with PE suspicion was the multidetector pulmonary CT angiography using 80 slice multi-detector CT (Prime SP Aquileon, Canon Medical Systems) after intravenous injection of 70 ml iodinated contrast agent (Iomeron 400 Mg I/mL) at a flow rate of 4 mL/s, followed by a 25 mL saline flush. The automatic bolus-tracking technique had the region of interest positioned at the level of the main pulmonary artery with a trigger threshold of 120 HU. CT scan settings were 120 kVp, 1 mm section thickness, 0.5mm interval, CTDIvol 4 mGy.

Two radiologist trainees with 2–4 years of experiences (S.A and R.A.) reviewed all images independently blinded to the clinical information, supervised by a senior radiologist with more than 10 years of experience (M.M.G). Each of the five lung lobes was assessed for percentage of the lobar involvement and classified as none (0%), minimal (1–25%), mild (26–50%), moderate (51–75%), or severe (76–100%), with corresponded score as 0, 1, 2, 3, or 4. The CT Total Severity Score (TSS) was reached by summing the five lobe scores (range from 0 to 20) (9).

The images were interpreted using the lung and mediastinum window setting. The CT images were assessed, following a standardized protocol, for the presence and distribution of the following abnormalities:

- Ground-glass opacities (GGO), defined as hazy areas of increased attenuation without obscuration of the underlying vascular markings.
- Interlobular septal thickening, intralobular septal line
- Crazy paving
- Consolidations, as parenchymal opacities obscuring underlying vessels
- Other no typical findings for COVID-19 pneumonia were recorded: pleural effusion, centrilobular, perilymphatic or random distributed nodules, tree in bud, etc.

We considered a compatible COVID-19 pneumonia if multilobar or patchy GGO, with or without interlobular septal thickening (crazy paving) or consolidation was present.

Whenever a chest X-ray was available and performed during the episode was recorded and analyzed.

### Ultrasound data collection

An emergency physician (Y.T.C) with long-standing experience in LUS (experienced sonologists on the basis of the American College of Emergency Physicians ultrasonographic guidelines and more than 10 ultrasound exams performed per week, 5 years of experience in performing and interpreting POCUS (6)) performed all ultrasound exams.

Participants were subjected to a LUS exam following a 12-zone protocol (10) (Figure 1). Each intercostal space of upper and lower parts of the anterior, lateral, and posterior regions of the left and right chest wall were carefully examined, and findings (pleural effusion, confluent and isolated B-lines, irregular pleural line, consolidations) were recorded (11-12). For each of the 12-zone a score from 0 to 3 was given according to the finding: irregular or isolated B-lines (1 point), confluent B-lines (2 points) and consolidations or pleural effusion (3 points). The total “Lung Ultrasonography (LUS) Score” was calculated by summing the scores of all 12 zones (range of possible scores, 0–36).

**Figure 1.**
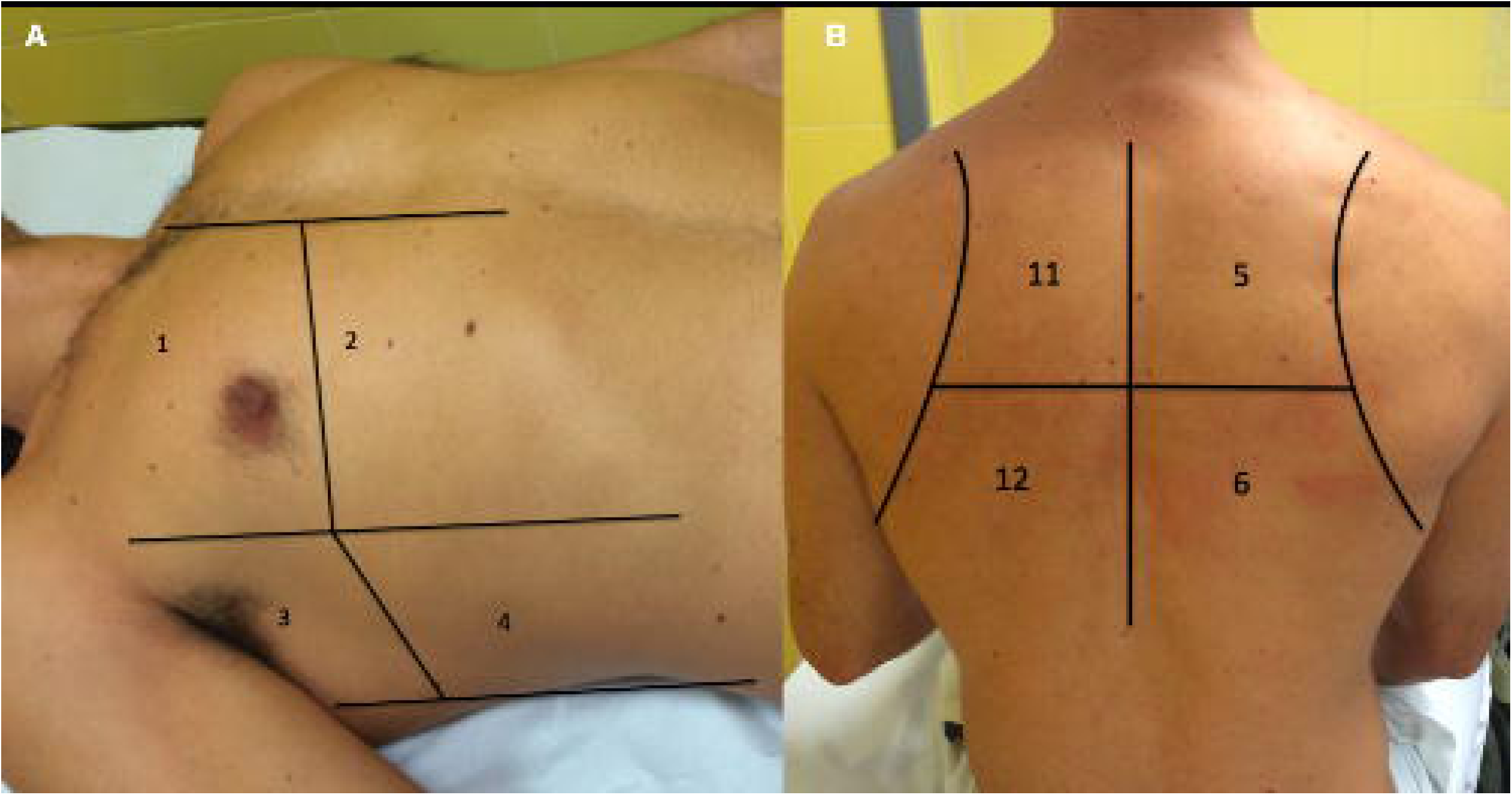
Representation of the 12-zone on chest. A: 1 and 2 right anterior, 3 and 4 right lateral, 5 and 6 left anterior (not represented), 7 and 8 left lateral (not represented). B: 5 and 6 right posterior, 11 and 12 left posterior.

A compatible LUS exam was considered a bilateral pattern of B-lines, isolated or confluent, irregular pleural line and subpleural consolidations.

The examinations were performed using a Butterfly IQ, a hand-held ultrasound system fitted with a curvilinear array transducer (1.5–4.5 MHz).

The physician was blinded to the patient past medical history, vital signs, symptoms, laboratory measurements and CT scan result.

### Outcome measures and definitions

The main purpose of this study is to evaluate the correlation between LUS findings and chest CT in COVID-19 patients (Figure 2 and 3).

**Figure 2.**
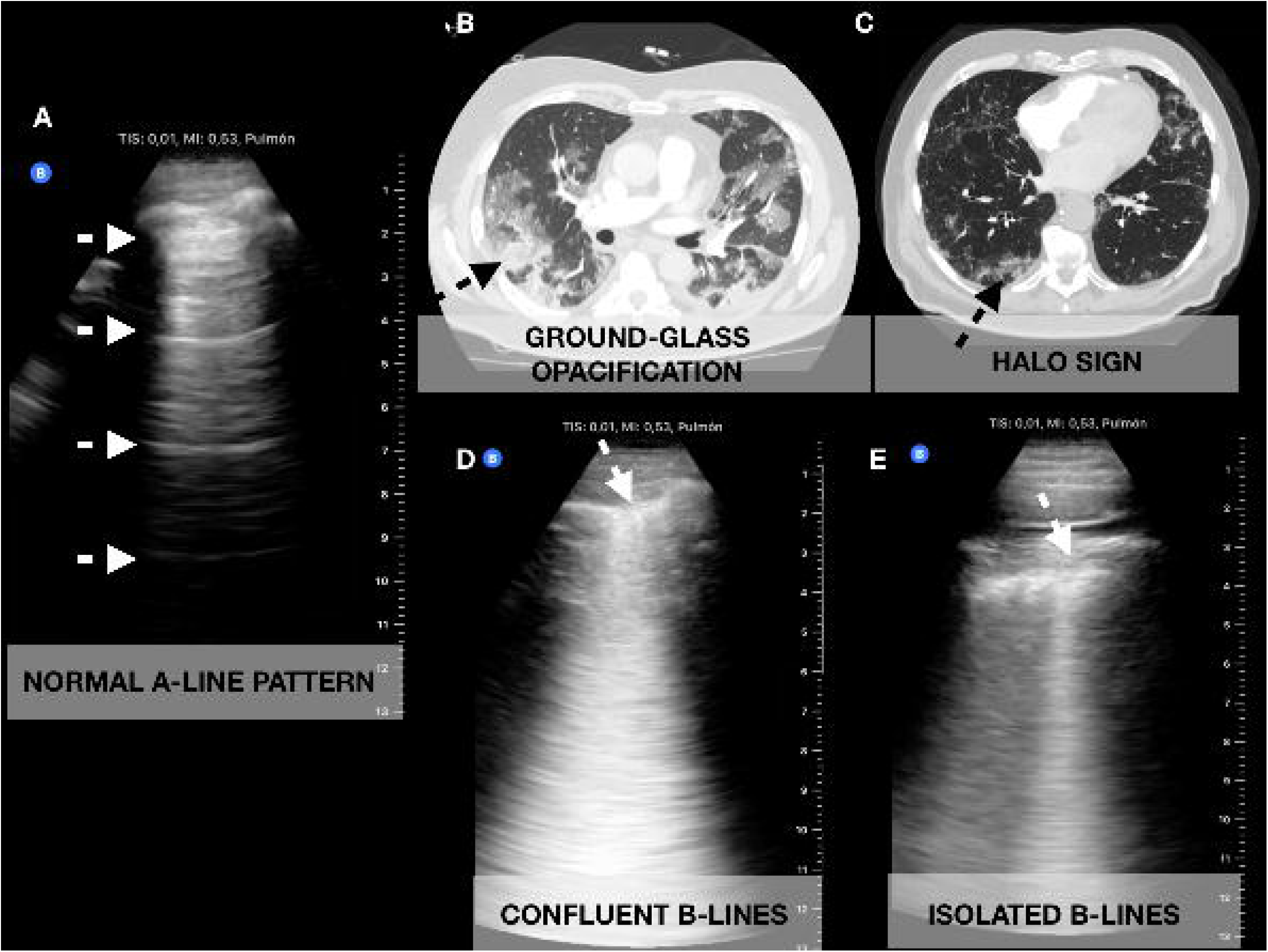
Correlation of chest Computed Tomography (CT) with Lung Ultrasonography (LUS) images obtained with a curvilinear probe. A) Normal A-line pattern on LUS. B) ground-glass opacification correlating with D) confluent B-lines, C) halo sign correlating with E) isolated B-lines.

**Figure 3.**
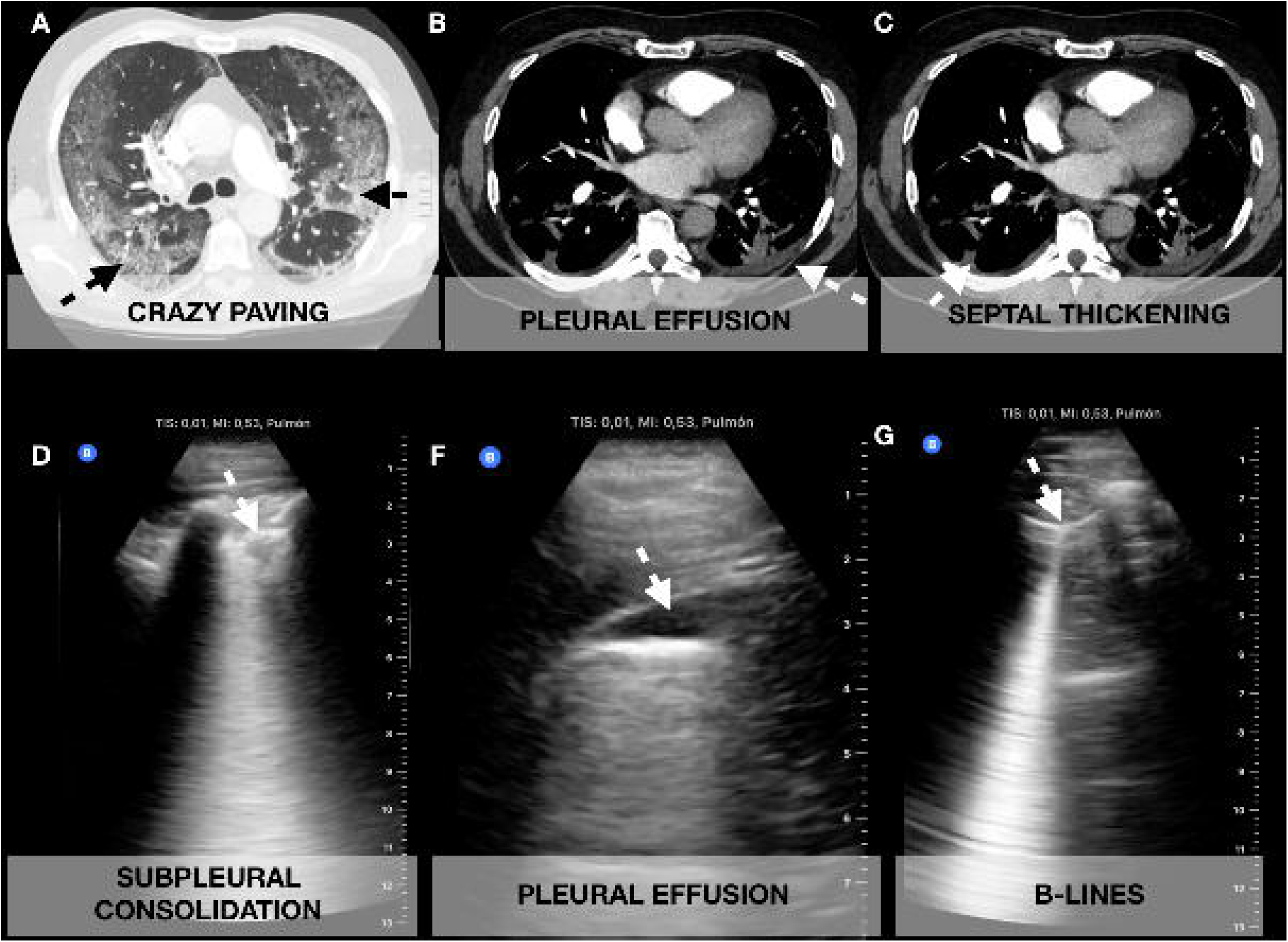
Correlation of chest Computed Tomography (CT) with Lung Ultrasonography (LUS) images obtained with a curvilinear probe. A) crazy paving correlating with D) subpleural consolidation, B) small pleural effusion seen in CT and in F) LUS. C) Septal thickening correlating with G) isolated B-lines.

We defined a confirmed case any patient with positive RT-PCR test, and clinically highly suspicious, any patient with dyspnea, fever, myasthenia, gastrointestinal symptoms, dry cough, ageusia or anosmia but negative RT-PCR.

### Statistical analysis

Baseline characteristics are presented as mean and standard deviation (SD) for continuous variables and count and proportions for categorical variables. The quantitative parameters were compared using a Mann-Whitney test for continuous variables and the χ^2^ or Fisher exact test for categorical one.

The intraclass correlation coefficients (ICC) was used to assess the degree of agreement between LUS Score and CT TSS. An ICC of less than 0.50 was considered poor, from 0.50 to 0.75 moderate, 0.75 to 0.90 was considered good and 0.90 to 1 excellent. The diagnostic performance of LUS compared to RT-PCR test to detect CT scan abnormalities was evaluated through a receiver operating characteristic curve analysis.

The correlations between continuous variables were tested using Spearman’s rho test for categorical variables. The sample size for correlation was calculated to detect a 20% of difference between LUS and CT findings, assuming a 95% confidence interval (CI) and power of 80%.

Mean values were reported along with 95% confidence intervals. Statistical significance was set at p value < 0.05.

Statistical analyses were conducted with IBM SPSS software v20.0 (SPSS Inc., Chicago, IL, USA).

## RESULTS

Fifty-one patients were consecutively enrolled between March and April 2020 (summarized in Table 1). The mean age was 61.4 years (Standard Deviation - SD 17.7). At the end of the first week follow-up, 34 patients were admitted to the hospital (66.7%), 4 were admitted to an intensive care unit (ICU, 7.8%), 6 patients had died (11.8%) and 17 were discharged to home (33.3%).

**Table 1.** Demographics and clinical characteristics of patients included on presentation (N=51).

| **Demographics** |  |
| --- | --- |
| Gender (female) - N (%) | 23 (45.1) |
| Age (years) mean (SD) | 61.4 (17.7) |
| **Past Medical History** |  |
| Cardiovascular disease - N (%) | 14 (27.5) |
| Pulmonary disease - N (%) | 12 (23.5) |
| Diabetes Mellitus - N (%) | 10 (19.6) |
| Chronic Kidney Disease - N (%) | 6 (11.8) |
| Immunosuppression - N (%) | 8 (15.8) |
| Hypertension - N (%) | 20 (39.2) |
| Malignancy - N (%) | 13 (25.5) |
| **Symptoms** |  |
| Dyspnea - N (%) | 29 (56.9) |
| Fever - N (%) | 23 (45.1) |
| Myasthenia - N (%) | 22 (43.1) |
| Gastrointestinal symptom - N (%) | 10 (19.6) |
| Cough - N (%) | 22 (43.1) |
| Ageusia/Anosmia - (N%) | 4 (7.8) |
| Onset of symptoms (days) mean (SD) | 3.5 (5.6) |
| **Physical exam** |  |
| SBP (mmHg) mean (SD) | 123.8 (18.5) |
| DBP (mmHg) mean (SD) | 72.8 (13.1) |
| Heart rate (bpm) mean (SD) | 94.9 (17.3) |
| Temperature (°C) mean (SD) | 36.5 (1.1) |
| SO2 (%) mean (SD) | 93 (5) |
| Respiratory rate (rpm) mean (SD) | 14.3 (4.1) |
| **Laboratory results - Mean (SD)** |  |
| WBC x 10<sup>9</sup>/L (SD) | 7.22 (3.3) |
| Lymphocyte x 10<sup>9</sup>/L (SD) | 1.27 (0.8) |
| Creatinine - mg/dL (SD) | 0.93 (0.49) |
| Urea - mg/dL (SD) | 45.2 (25.6) |
| ALT - U/L (SD) | 57.8 (128.5) |
| LDH - U/L (SD) | 382.5 (291.9) |
| D-dimer - ng/mL (SD) | 6870.9 (14324) |
| C-Reactive Protein - mg/dL (SD) | 72.0 (103.1) |
| Troponin I - ng/mL (SD) | 296.4 (1285.3) |
| NT-proBNP - pg/mL (SD) | 2963.0 (2837.9) |
| IL-6 - pg/mL (SD) | 214.5 (351.8) |
| Ferritin - ng/mL (SD) | 873.6 (1567.5) |
| ***SARS-CoV-2 (RT-PCR) test*** | **48 (94.1)** |
| Positive - N (%) | 23 (47.9) |
| Negative - N (%) | 23 (47.9) |
| Indeterminate - N (%) | 2 (4.2) |
| ***Follow-up*** |  |
| Admission - N (%) | 34 (66.7) |
| ICU - N (%) | 4 (7.8) |
| Discharge - N (%) | 17 (33.3) |
| Mortality - N (%) | 6 (11.8) |
| DBP: diastolic blood pressure; IL-6: Interleukin 6; LDH: Lactate dehydrogenase; NT-ProBNP: N-terminal pro-brain natriuretic peptide; PCT: procalcitonin; RT-PCR: reverse transcriptase polymerase chain reaction; SARS-CoV-2: Severe acute respiratory syndrome coronavirus 2; SBP: systolic blood pressure; SD: standard deviation. |  |

**Table 2.**
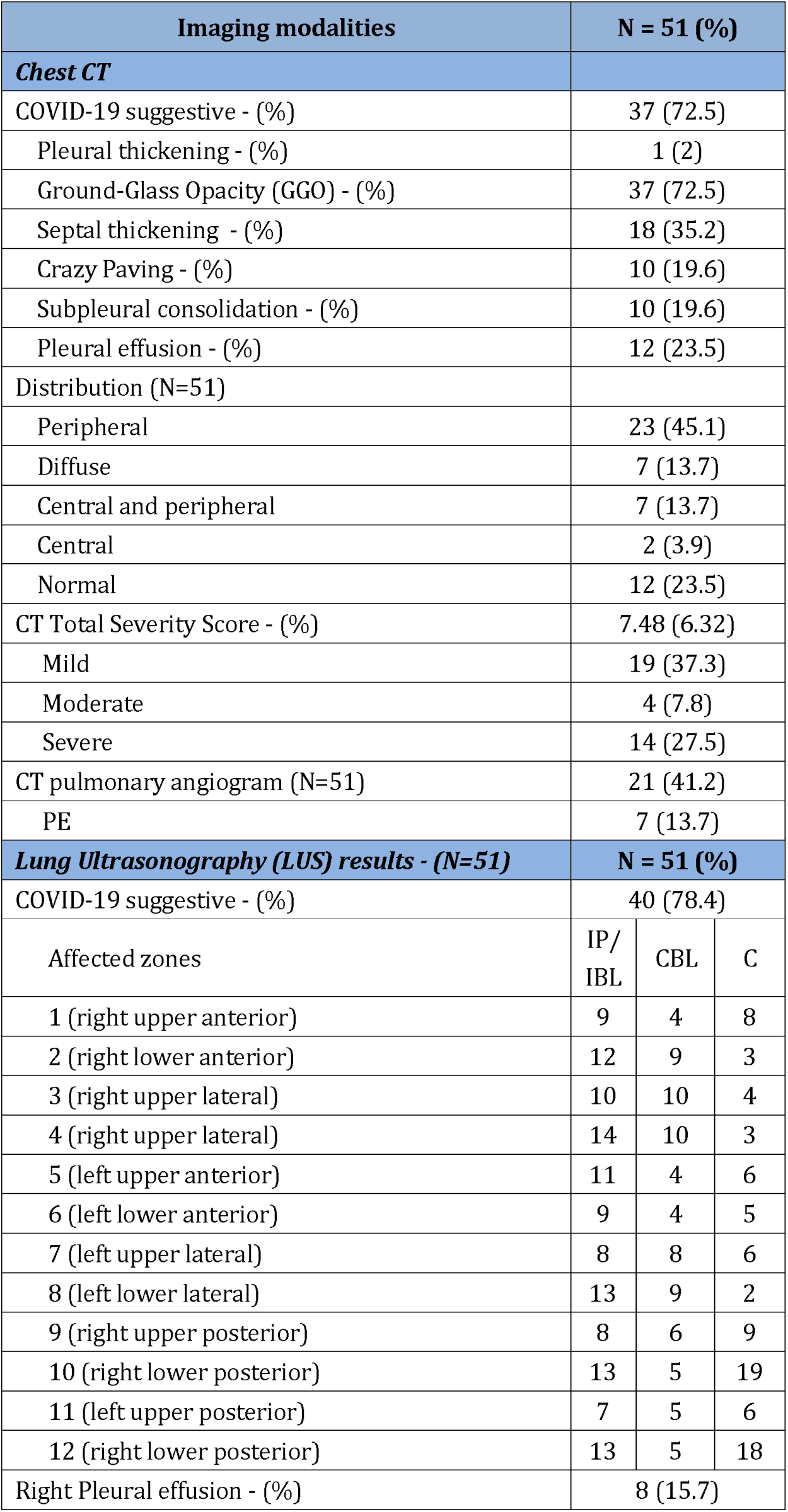
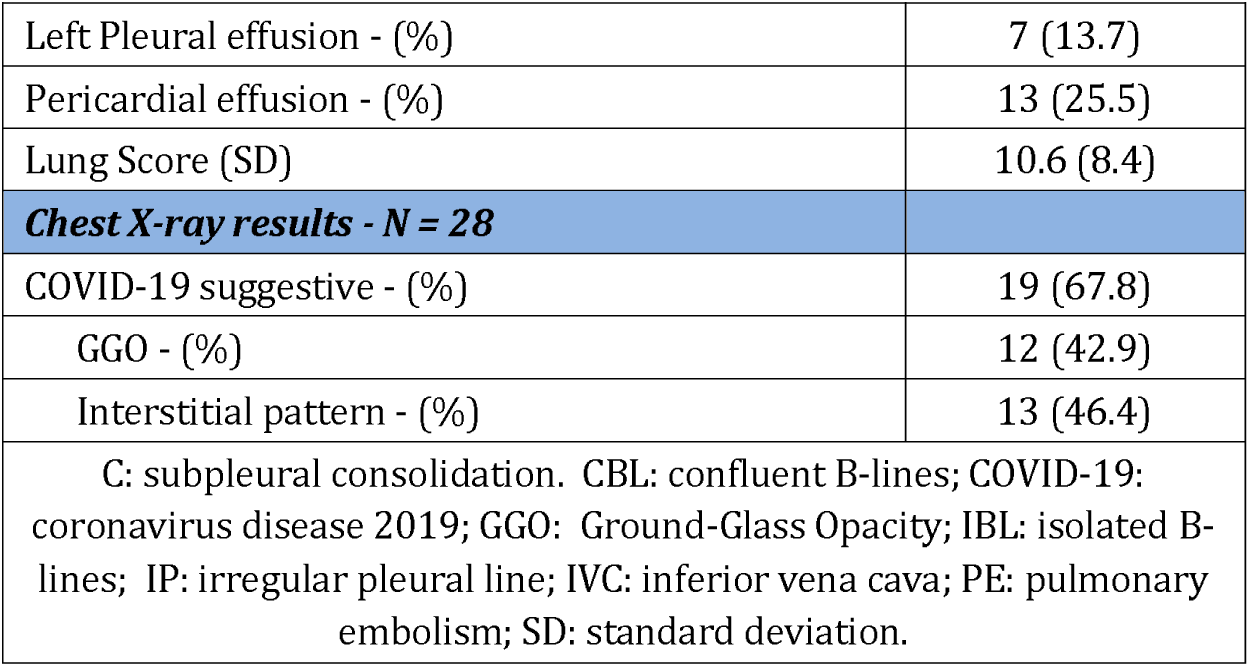
Imaging modalities (chest CT, Lung Ultrasonography and chest X-ray) findings of patients included.

| Imaging modalities | N = 51 (%) |  |  |
| --- | --- | --- | --- |
| ***Chest CT*** |  |  |  |
| COVID-19 suggestive - (%) | 37 (72.5) |  |  |
| Pleural thickening - (%) | 1 (2) |  |  |
| Ground-Glass Opacity (GGO) - (%) | 37 (72.5) |  |  |
| Septal thickening - (%) | 18 (35.2) |  |  |
| Crazy Paving - (%) | 10 (19.6) |  |  |
| Subpleural consolidation - (%) | 10 (19.6) |  |  |
| Pleural effusion - (%) | 12 (23.5) |  |  |
| Distribution (N=51) |  |  |  |
| Peripheral | 23 (45.1) |  |  |
| Diffuse | 7 (13.7) |  |  |
| Central and peripheral | 7 (13.7) |  |  |
| Central | 2 (3.9) |  |  |
| Normal | 12 (23.5) |  |  |
| CT Total Severity Score - (%) | 7.48 (6.32) |  |  |
| Mild | 19 (37.3) |  |  |
| Moderate | 4 (7.8) |  |  |
| Severe | 14 (27.5) |  |  |
| CT pulmonary angiogram (N=51) | 21 (41.2) |  |  |
| PE | 7 (13.7) |  |  |
| ***Lung Ultrasonography (LUS) results - (N=51)*** | **N = 51 (%)** |  |  |
| COVID-19 suggestive - (%) | 40 (78.4) |  |  |
| Affected zones | IP/<br>IBL | CBL | C |
| 1 (right upper anterior) | 9 | 4 | 8 |
| 2 (right lower anterior) | 12 | 9 | 3 |
| 3 (right upper lateral) | 10 | 10 | 4 |
| 4 (right upper lateral) | 14 | 10 | 3 |
| 5 (left upper anterior) | 11 | 4 | 6 |
| 6 (left lower anterior) | 9 | 4 | 5 |
| 7 (left upper lateral) | 8 | 8 | 6 |
| 8 (left lower lateral) | 13 | 9 | 2 |
| 9 (right upper posterior) | 8 | 6 | 9 |
| 10 (right lower posterior) | 13 | 5 | 19 |
| 11 (left upper posterior) | 7 | 5 | 6 |
| 12 (right lower posterior) | 13 | 5 | 18 |
| Right Pleural effusion - (%) | 8 (15.7) |  |  |

|  |  |
| --- | --- |
| Left Pleural effusion - (%) | 7 (13.7) |
| Pericardial effusion - (%) | 13 (25.5) |
| Lung Score (SD) | 10.6 (8.4) |
| ***Chest X-ray results - N = 28*** |  |
| COVID-19 suggestive - (%) | 19 (67.8) |
| GGO - (%) | 12 (42.9) |
| Interstitial pattern - (%) | 13 (46.4) |
| C: subpleural consolidation. CBL: confluent B-lines; COVID-19: coronavirus disease 2019; GGO: Ground-Glass Opacity; IBL: isolated B-lines; IP: irregular pleural line; IVC: inferior vena cava; PE: pulmonary embolism; SD: standard deviation. |  |

Approximately half of the patients (54.9%) had a chest X-ray, in whom 33.2% were normal.

Ground-glass opacities (GGO) were present in 37 patients (72.5%), with peripheral or diffuse involvement, followed by septal thickening (18 patients, 35.2%).

There were two patients with only central involvement on CT, one patient had a mild cardiac failure and another patient had viral bronchiolitis.

The most common finding and affected zones on LUS were subpleural consolidations on the posterior lower lobes. The mean LUS score was 10.6 (SD 8.4). The mean CT TSS was 7.48 (SD 6.32). **The LUS Score had a good correlation with CT TSS** (Intraclass Correlation Coefficient - ICC 0.803, 95% Confidence Interval - CI 0.601-0.903, *p* < 0.001). The age showed a moderate correlation to LUS Score (rho = 0.486; p < 0.001) but not with CT TSS (p = 0.247). Oxygen saturation (SO2) correlated stronger to LUS Score (rho = -0.553; p < 0.001) than to CT TSS (rho = - 0.360; p = 0.043). Respiratory rate correlated stronger to LUS Score (rho = 0.529; p < 0.001) than to CT TSS (rho = 0.429; p = 0.020). As well, inflammatory markers such as C-reactive protein (CRP) correlated better with LUS Score (rho = 0.600; p = 0.004) than to CT TSS (rho = 0.479; p = 0.005).

Radiological signs suggestive or highly compatible with COVID-19 were present in thirty-seven patients (72.5%) on CT scan and forty (78.4%) in LUS exam. All thirty-seven patients with abnormal findings on CT were correctly diagnosed with LUS (Odds Ratio - OR: 13.333, 95% CI: 4.490-39.591, *p* < 0.001) with a sensitivity (S) of 100.0% and a specificity (Sp) of 78.6%, positive predictive value (PPV) of 92.5% and negative (NPV) of 100.0%.

Although, there were three patients with LUS findings compatible of COVID-19, on CT two were informed as having a viral bronchiolitis and one had pulmonary metastatic disease.

An analysis of receiver operating characteristic curves (Figure 4) showed that the area under the curve [AUC] for LUS (86.4%; 95% CI, 70.2%–100%, p<0.001) was better in comparison to RT-PCR (63.4%; 95% CI, 45.0%–81.8%; p=0.181) for detection of CT abnormalities.

**Figure 4.**
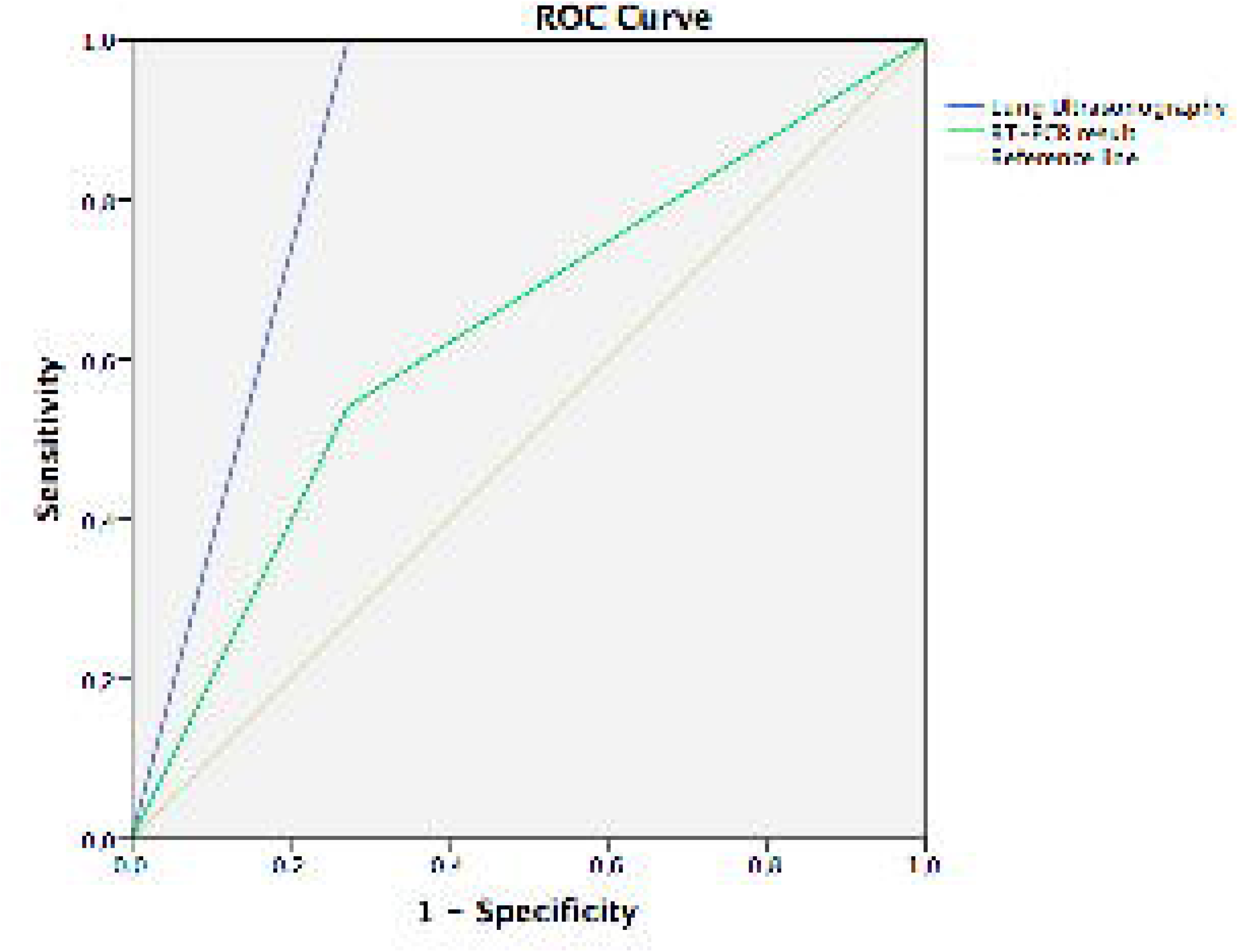
Receiver Operator Characteristic (ROC) curve for RT-PCR test, AUC of 63.4% and Lung Ultrasonography (LUS) exam, area under the curve (AUC) of 86.4% for detecting CT abnormalities.

Therefore, there was no missed diagnosis of COVID-19 with LUS compared to CT in our cohort.

## DISCUSSION

In our study, we found an excellent correlation between CT and LUS. All abnormal CT findings were detected on LUS, therefore no abnormal CT findings were labeled as normal on LUS. In other words, with this technic the proportion of false negative rates will be really low, which in this pandemic is a key question, in order to avoid additional infections.

There is growing literature regarding the diagnostic challenges (4-5) in COVID-19 patients. **The positivity** rate of RT-PCR has been quantified as 63% in nasal swab and 32% in pharyngeal swab (13), similar to our results, we found a positive rate of only 47.9%. Due to its limitations, diagnostic imaging plays a key role in the management of these patients.

A study of 1049 patients undergoing chest CT scan and RT-PCR testing determined that CT abnormalities had a highly sensitivity for diagnosis of COVID-19 patients (1), suggesting that CT scan should be considered as a screening tool, especially in epidemic areas with high pre-test probability. Therefore in many centers CT scans have been replaced for chest X-ray. However, the use of CT scan in the ED has many limitations, such as the radiation exposure, especially for mild illness, the low availability and the contraindication of its use in unstable patients. Also, we found a proportion of normal chest CT that is relatively high (27.4%), but similar to those previously reported (30.8%) (9).

Preliminary reports in COVID-19 era, suggest there is a correlation of LUS findings to those of the CT scan (14-15). These reports have characterized LUS findings in COVID-19 patients. Moreover, Soldati et al (16) have proposed a standardized approach to performing LUS in these patients, including a 14-zone technique, and a scoring system to quantify severity of lung involvement. Although we agree there should be a consensus in the LUS exam method, the 12-zone is more extended and validated (12).

There have been reports regarding the cardiac injury in COVID-19 patients that ranges from 7.2%-14% (17-18). In our study, we found one young patient that had a normal LUS exam, which prompted a sonographic focused cardiovascular assessment, showing a left ventricular dysfunction and pericardial effusion, allowing immediate therapy adjustment. His chest CT scan was unremarkable.

There were 2 patients with absence of typical findings for COVID-19 (central distribution); one patient had acute bronchiolitis and the other patient a decompensated heart failure episode. Both of them, although had mainly central involvement on CT, had also some degree of peripheral involvement of the lungs, which was seen on LUS, misidentifying as COVID-19 suspicious findings. This is one of the main limitations of LUS (19), the low specificity, since the findings might overlap with those from other lung disease etiologies, such as other viral illnesses, pulmonary infarction or metastatic disease. Although, the same might happen with CT scans, that can misidentify other viral pneumonias as COVID-19. However, in this pandemic, positive LUS or CT features, even in negative RT-PCR can still be highly suggestive of COVID-19 infection.

There are several advantages of performing LUS over CT scan, particularly when performed with portable handheld ultrasound devices, may provide an inexpensive, accessible, portable, user-friendly, and easy-to-disinfect method for assessing progression of cardiopulmonary pathology in patients with COVID-19 pneumonia. Moreover, avoids having to transport a patient with suspected COVID-19 to radiology (exposing other patients or health care providers).

In our study we found a high correlation between LUS findings and chest CT abnormalities suggestive of lung involvement due to SARS-CoV-2 infection. As previously reported (5), most of the patients had a predominance of peripheral involvement of both lungs (92.5%, 37 out of 40 patients with abnormal CT findings), which can be reached with ultrasound. Notably, the LUS score showed a better correlation than CT TSS in demographic data (age), physical exam (respiratory rate, SO2) and other established biomarkers (CRP) with proven utility in this setting, this higher correlation should interpreted cautiously, although might suggest that a 12-zone LUS Score, with the representation of posterior lobes in at least half of the zones (in comparison to a 5 lobes division, where posterior lobes represent 2) could better reflect the physiological state of the patient.

Therefore this technique could be more easily replaced with LUS as it would be more accessible during the pandemic, especially accelerated as Artificial Intelligence algorithms to easy recognize COVID-19 related pathology and telemedicine programs are developed.

### - Strengths

To our knowledge, this is the first study evaluating the correlation of LUS with CT scan, with the diagnostic and prognostic implications. We evaluated the radiologic burden (CT and LUS) with clinical manifestation, laboratory results and outcomes.

### - Limitations

There are several limitations to consider. The main limitation is the small sample. Another limitation is that selection bias might have occurred. The expert sonographer performed all ultrasound scans on a consecutive sample selected based on his availability (during his working hours), which limits the generalizability of our results. This was mitigated by the variable schedule and changing shifts, unpredictable a priori (in continuous care). Additionally, false negative ultrasound or CT might be found in the initial stage of the disease, before lung involvement, consequently imaging techniques should be considered a complement to RT-PCR and laboratory tests.

The study was not powered to evaluate the performance of a diagnostic strategy based on LUS exam; therefore, for this purpose, the study can only be considered hypothesis generating. Thus, the results from this study open an opportunity to further investigate the use of ultrasound in different settings and clinical scenarios, especially in the follow-up.

We want to share our study findings, given the urgent need for different strategies in order to better manage COVID-19 patients, and diminish the SARS-CoV-2 spread and its prognosis in the current pandemic context. As the shortage of resources constitutes an undeniable public health threat, we consider LUS to be a potential solution, and recommend that it should be performed as a first-line and follow-up imaging test for COVID-19 patients.

## CONCLUSIONS

**LUS presents similar accuracy compared to chest CT to detect lung abnormalities in COVID-19 patients**. In this pandemic, as the shortage of resources constitutes an undeniable public health threat, LUS can play a strategic role that has the potential to impact the management of these patients.

## Data Availability

yes, on demand

## Acknowledgments

- The authors have declared no conflicts of interest.

